# Applications of Data Characteristic AI-assisted Raman Spectroscopy in Pathological Classification

**DOI:** 10.1101/2023.09.05.23295054

**Authors:** Xun Chen, Jianghao Shen, Chang Liu, Xiaoyu Shi, Weichen Feng, Hongyi Sun, Weifeng Zhang, Shengpai Zhang, Yuqing Jiao, Jing Chen, Kun Hao, Qi Gao, Yitong Li, Weili Hong, Pu Wang, Limin Feng, Shuhua Yue

## Abstract

Raman spectroscopy has been widely used for label-free biomolecular analysis of cell and tissue for pathological diagnosis *in vitro* and *in vivo*. AI technology facilitates disease diagnosis based on Raman spectroscopy including machine learning (PCA and SVM), manifold learning (UMAP) and deep learning (ResNet and AlexNet). However, it is not clear how to optimize the appropriate AI classification model for different types of Raman spectral data. Here, We selected five representative Raman spectral datasets, including endometrial carcinoma, hepatoma extracellular vesicles, bacteria, melanoma cell, diabetic skin, with different characteristics regarding sample size, spectral data size, Raman shift range, tissue sites, Kullback-Leibler (KL) divergence, and key Raman shifts, explore the performance of different AI models (e.g. PCA-SVM, SVM, UMAP-SVM, ResNet or AlexNet). Tissue sites mean that spectral collection sites from sample, KL divergence means the divergence between spectra of different types. We found that for dataset of large spectral data size, Resnet performed better than PCA-SVM and UMAP, for dataset of small spectral data size, PCA-SVM or UMAP performed better. We also optimized the network parameters (e.g. principal components, activation function, and loss function) of AI model based on data characteristics. Using AI classification models, the mean area under receiver operating characteristic curves (AUC) for representative datasets reached 0.966, with mean sensitivity of 89.6%, mean specificity of 95.4%, mean accuracy of 93.4%, and mean time expense of 5 seconds. By using data characteristic assisted AI classification model, the accuracy improve from 85.1% to 94.6% for endometrial carcinoma grading, from 77.l% to 90.7% for hepatoma extracellular vesicles detection, from 89.3% to 99.7% for melanoma cell detection, from 88.1% to 97.9% for bacterial identification, from 53.7% to 85.5% for diabetic skin screening. Furthermore, according to the saliency maps, we found classification-associated biomolecules (e.g. nucleic acid, tyrosine, tryptophan, cholesteryl ester, fatty acid, and collagen), which contribute to the pathological diagnosis classification. Data characteristic assisted AI classification model was demonstrated to improve the robustness and accuracy of Raman spectroscopy in pathological classification. Collectively, this study opens up new opportunities for accurate and rapid Raman optical biopsy.

## INTRODUCTION

The vibrational modes of molecules provide an intrinsic contrast mechanism for detecting compositions in biological system. Raman scattering enables in vitro and in vivo characterization as a sensitive probe of chemical composition. In the past 30 years, Raman spectroscopy has been widely used for molecular analysis of biological samples^1,2^. Advances in setup, methodology, and data analysis enable excellent prospects for a wide range of laboratory and clinical uses.

However, Raman signal is intrinsically composed of overlapping and broad features, which make it hard to read for pathologists and doctors. For example, Raman spectra from normal and tumor tissues generally are similar, and spectral difference cannot be distinguished accurately. To observe the subtle spectral difference, several spectral data analysis algorithms have been reported to enable the classification of spectra from samples, for instance, bacterial identification^3^, pathological diagnosis^4^, and treatment response^5^ etc. The analysis efficiency is highly depending on the analysis algorithm and diagnostic models. However, the robustness of conventional models is not strong enough. Due to the diversity and heterogeneity of the biological system, the prediction accuracy will be substantially reduced when the model is applied to the extra dataset acquired. AI models integrated the chemical information within Raman spectra, which were potential for accurate and robust classification.

For instance, machine intelligent methods (machine learning, manifold learning and deep learning) were developed to improve the accuracy and robustness of spectral classification by Raman spectroscopy^6,7^. Machine learning (ML) models such as principal component analysis (PCA), linear discriminant analysis (LDA), support vector machine learning (SVM) and logistic regression (LG) etc. have been demonstrated to differentiate Raman spectra^8–11^. Manifold learning models such as uniform manifold approximation and projection (UMAP) were also used to process Raman spectra with nonlinear dimensional reduction^12^. It is possible to model Raman spectra with such a topological characteristic using UMAP. The embedding in UMAP was demonstrated to differentiate fibroblasts and iPSC^12^.

Moreover, deep learning (DL) method such as convolutional neural networks based deep learning algorithms^13–16^ have also been used to classify Raman spectra. Huang et al. developed a Raman-specified convolutional neural networks, which performed better than ML models, for diagnosis of nasopharyngeal carcinoma and assessment of post-treatment efficacy^5^. Raman spectroscopy combined with a long short-term memory (LSTM) was developed to improve the accuracy of the identification of marine pathogens^17^.

To figure out the contribution of Raman shifts in classification, Huang et al. firstly used t-test method and found that Raman shift related to collagen, protein, and nuclei acid contribute to tumor malignant progression^18^. Lin et al. then used PCA-LDA method and analyzed the contribution of Raman shifts by PCA components^19^. In another study, Erzina et al. added the binary stochastic filtering (BSF) layers to the classifier after each of the CNN inputs to quantify the molecular contribution^20^.

However, it is not clear how to optimize the appropriate AI classification model for different types of Raman spectral data. The development procedures may waste a lot of time to find best models and adjust model parameters. Here, our hypothesis is that the parameter size of model, which is commonly considered to be related to the fitting capability, of the best model will increase with larger spectral size and less spectral divergence. As shown in **Figure 1(a)**, we used sample size, spectral data size, Raman shift range, tissue sites, KL divergence, and key Raman shifts as the indicators to evaluate the characteristics of each dataset. For representative datasets, the performance of diagnostic models between deep learning, machine learning and UMAP were shown in **table S1** and **Figure 1(b). Figure 1(c)** It was a positive correlation between the best model parameter size with the spectral data size instead of merely tissue sites/sample spots. We also observed a positive correlation between parameter size and Raman shift range.

**Figure 1.**
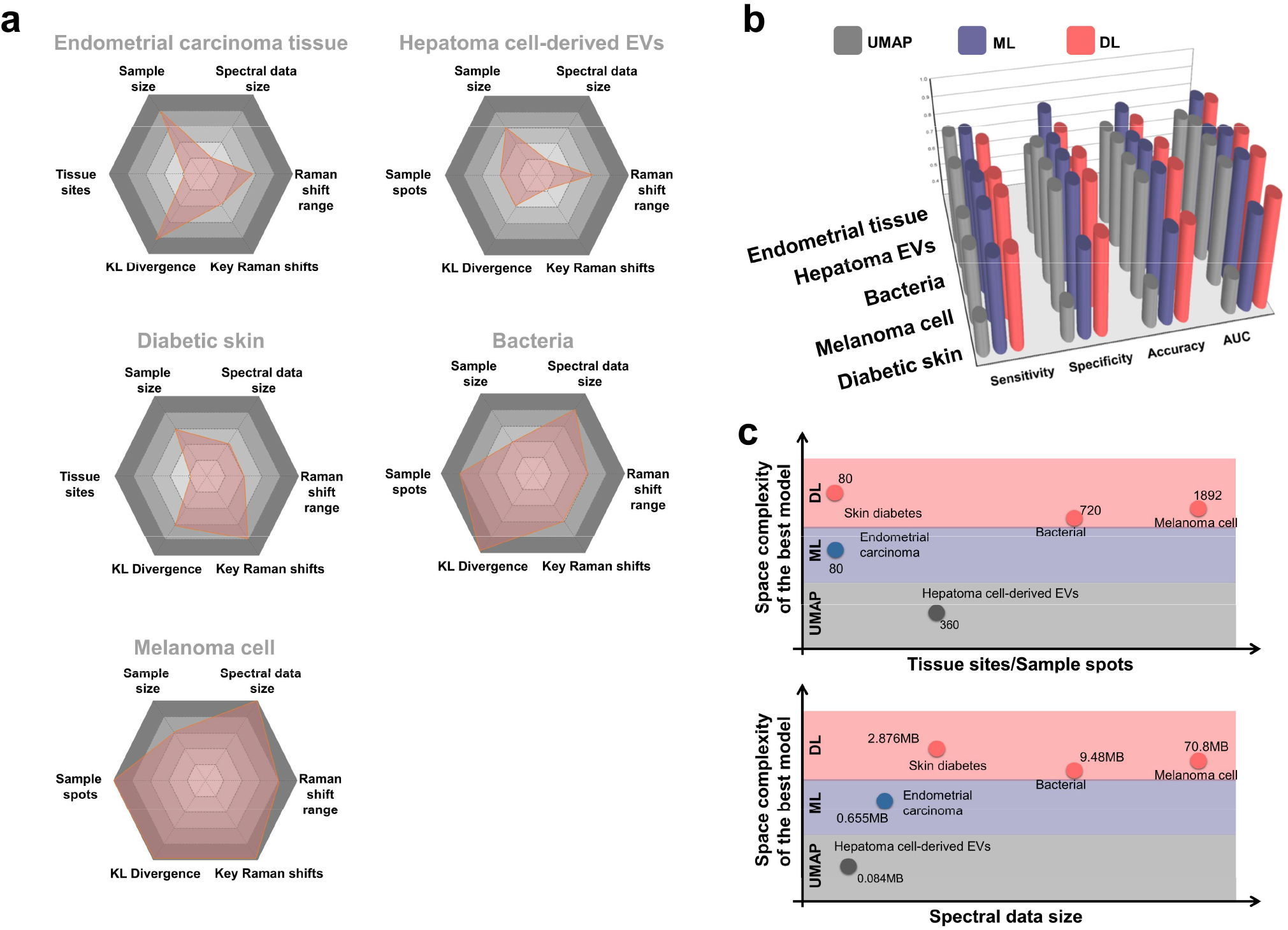
Raman spectrum characterizations of representative datasets and data characteristic input spectra versus best AI classification models. We used five representative datasets including endometrial carcinoma, hepatoma cell EVs, bacteria, melanoma cell, and diabetic skin. (a) Sample size, spectral data size, sample spots/tissue sites, Raman shift range, KL divergence, and key Raman shift. (b) Sensitivity, specificity, accuracy and AUCs of five Raman dataset by comparing DL, ML, and UMAP models (c) Best model parameter size (.*h5 format*) versus either sample spots/tissue sites or spectral data size (.*npy format*) respectively.

Furthermore, we analyzed molecular contribution of the best model with Raman shift explanation, and especially we proposed a new method to calculate class weight using UMAP. We also used the BSF in DL method to analyze the contribution of Raman spectra and found the contribution molecules such as glucose, collagen and protein, nucleic acids, saturated and unsaturated fatty acid and lipids etc. in representative datasets. All these improvements may benefit the robustness of data characteristic AI-classification model and put Raman spectroscopy into rapid pathological classification.

## METHODS AND EXPERIMENTS

We used five Raman dataset including three data we collected using our setup, and two public data from previous papers. For our collection data, two spontaneous Raman data were collected from endometrial cancer and brain cancer tissues using Raman probe-based system for intraoperative pathological diagnosis. Another SERS was collected for EVs detection using the same system. Another Raman data was collected for bacterial identification using high-numerical aperture (NA) Raman confocal microscopy. For public data, one is that Raman spectra from ear lobe, inner arm thumb nail, and median cubital vein could screen diabetes mellitus with combining machine learning algorithm and the Raman probe tool^21^. Another is that SERS of normal and cancer cells medium with or without serum could be recognized via the combination of functionalized SERS surfaces and convolutional neural network with independent inputs^20^.

The Raman probe spectroscopy system which we used for endometrial cancer and GBM diagnosis, and melanoma cell detection is composed of Raman probe with filters (RamanProbe, Inphotonics Inc.), 785nm laser (o8NLDM, Cobolt Inc.) and high-sensitive spectrometer with ddpCCD (Acton 785, Princeton Instrumentation Inc.). The laser excitation power for the tissue Raman collection is 100mW, and the exposure time of single spectrum is 5-10 second. The numerical aperture (NA) of Raman probe (1cm in diameter) is 0.22.

The confocal Raman microscopy system which we used for bacterial identification is composed of a Raman spectrometer (KYMERA-328I-A, Andor) with a 707 nm laser source. The laser (tunable 700–990 nm wavelength, Applied Physics & Electronics Inc.) power at the sample was ∼10 mW after a 60× water objective (NA= 1.2), and the exposure time we acquired the single spectrum was 1 second. The grating was 300 l/mm.

The original spectral data contains various noise and auto-fluorescence background; therefore, the spectra need to be processed before being input into the deep learning model. The pre-processing takes four steps: (1) wavenumber selection; (2) background subtraction; (3) smoothing; (4) normalization. In brief, the wavenumber between 400-1800 cm-1 was selected as the region of interest. The asymmetric least-squares method was applied to subtract the background signal. The data were then smoothed by a Savitzky-Golay filter to reduce the noise and increase the signal-to-noise ratio. All the processing mentioned above was done by Python 3.7 scipy 1.8.0.

We calculated the Kullback–Leibler (KL) divergence^22^ and significant wave-number points from total wave-number points for each Raman and SERS dataset. The KL divergence formular between spectrum from different categories was below:

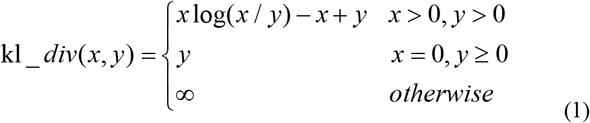

For instance, x is a spectrum of normal cell, and y is a spectrum of cancer cell. This process was done by Python 3.7 scipy 1.8.0. The significant wave-number points were calculated by using variance threshold^23^. The significant wave-number points are that the Raman shift with the variance which is larger than 0.01 after the value 0-1 normalization. This process was done by Python 3.7 sklearn 0.24.2.

We also developed a UMAP class weight method to calculate Raman shift contribution. The class weight was simulated by the schematic below:

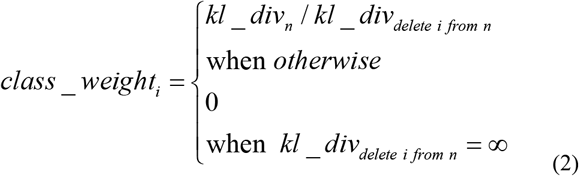

Among the formular, *i* is Raman shift and *n* is total Raman shift. We calculate each class weight of Raman shift of *kl_div*_*n*_ and *kl_div*_*delete i from n*_ after UMAP. Additionally, the class weight of PCA were simulated by feature importance coefficients using *Python 3*.*7 sklearn 0*.*24*.*2*. We got PCA components or UMAP components^24^ during the pre-process dimensional reduction by *Python 3*.*7 sklearn 0*.*24*.*2* and *UMAP-learn 0*.*5*.*3*. UMAP^25^ has no computational restrictions on embedding dimension, making it viable as a general purpose dimension reduction technique for machine learning. After UMAP and PCA pre-process of Raman data, we use SVM to build diagnosis model. The scheme of UMAP, PCA and SVM was shown in **Supplementary Note 1** and **Figure S1**. Especially, we developed a new feature selection based UMAP spectra analysis algorithm. The feature selection eliminated Raman shift with low variance and low feature importance, by the variance threshold and sequential feature selection (SFS) algorithms^23^.

The details of training set and networks of AlexNet and ResNet were described in the **Supplementary Note 1** and **Figure S2**. The training process of training loss and validation loss were shown in **Figure S3**.The class weight of deep learning models such as AlexNet and ResNet were simulated by the binary stochastic filtering (BSF) feature selection methods^26^. This was done by *Python 3*.*7 keras 2*.*2*.*4* and *tensorflow 1*.*14*.*0*.

Statistically significant differences were reported when p<0.05. Statistical analysis was performed using Origin (Origin Software, Inc). The multivariate classification for bacterial ID and melanoma cell detection was evaluated with a multiclass receiver operating characteristic (ROC) analysis^27^ according to the method described in this website^28^. By using trained AlexNet and ResNet, probabilities of bacteria and cell categories were predicted. A ROC curve was generated by continuously varying the threshold of the probability for each category based on the ground truth. The area under the ROC curve (AUC) ranging from 0 to 1 evaluates the ability of a model to accurately distinguish different categories. The details of multi-class confusion matrix and ROCs were described in **Figures S4** and **S5**.

## RESULTS

We selected five representative datasets from endometrial cancer tissue, hepatoma cell EVs, bacteria, melanoma cell, and diabetic skin based on data characteristics regarding sample size, spectral data size, Raman shift range, tissue sites, KL divergence, and key Raman shifts in the **Figure 1(a)**. Based on five Raman demo, data characteristics using hexagonal figures with each distribution type were summarized. For instance, melanoma cell dataset has more spectral data size, hepatoma cell EVs dataset has less spectral data size. Meanwhile, endometrial cancer tissue dataset has less tissue sites, diabetic skin dataset has more tissue sites. We focus on comparing the best model from DL, manifold learning and ML methods (PCA+SVM, SVM, UMAP+SVM and AlexNet, ResNet) with better AUCs, sensitivity, specificity and accuracy in the **Figure 1(b)**. The best AI classification model parameter size was related with either Raman spectral data size or spectral tissue sites as shown in **Figure 1(c)**. The details are described below.

### Endometrial Cancer Diagnosis

We demonstrate that the performances of spontaneous Raman classification by using machine learning, manifold learning and deep learning algorithms. The first representative dataset was from endometrial cancer tissues. The malignant level of endometrial cancer is highly related with the treatment strategies. There are some fertility-sparing treatments in patients with early endometrial cancer (EEC) or atypical complex hyperplasia (ACH)^29^. However, current diagnostic model for endometrial cancer was rarely studied by Raman spectroscopy. Here we aim to collect Raman database from endometrial tissues in-vitro firstly and differentiate the benign and malignant endometrial cancer. We built models (PCA+SVM, SVM, UMAP+SVM and AlexNet, ResNet) and differentiate the benign and malignant endometrial cancer tissues using our high-sensitivity Raman-probe spectroscopy system.

In **Figure 1(a)**, the divergence between benign and malignant is higher among five datasets, and the data size of Raman spectra from endometrial cancer is lower among five datasets, therefore simple ML based model may work better for endometrial cancer diagnosis. By comparing the AUCs of DL, manifold learning and ML methods, it indicated that PCA+SVM was the best. By PCA preprocessing, the principal components with higher variance were the input of SVM model. The AUC of PCA-SVM increased by 0.016 on average in 10 repeats compared with SVM.

The average spectra of benign and malignant are shown in **Figure 2(a)**. We compared the discriminant distribution between benign and malignant spectra using UMAP and PCA dimensional reduction methods in **Figure 2(b, and c)**. The visualization of UMAP is better than PCA pre-process, and UMAP components separate each other after the feature selection. By comparing the confusion matrix of four methods of UMAP+SVM, SVM, SVM+PCA and AlexNet in **Figure 2(d)**, we found that the best model in this case was PCA+SVM, and the total parameter size of these two models were 0.359 MB, with the AUC of 0.960±0.002 in the **table S1**. Additionally, we calculated the Raman shift class weight, as shown in **Figure 2(d)**. The top contribution molecules with corresponding Raman shift were (amide I band - (C=O) stretching mode of proteins, collagen) (∼1654 cm^-1^), stretching mode (C=C) tryptophan/porphyrin of protein (∼1548 cm^-1^ and 1615 cm^-1^), CH_2_ bending mode of proteins and lipids (∼1442 cm^-1^), saccharide (∼1368 cm^-1^), asymmetric stretch PO_2_^--^ nucleic acids (∼1221 cm^-1^)^8,18,30^.

**Figure 2.**
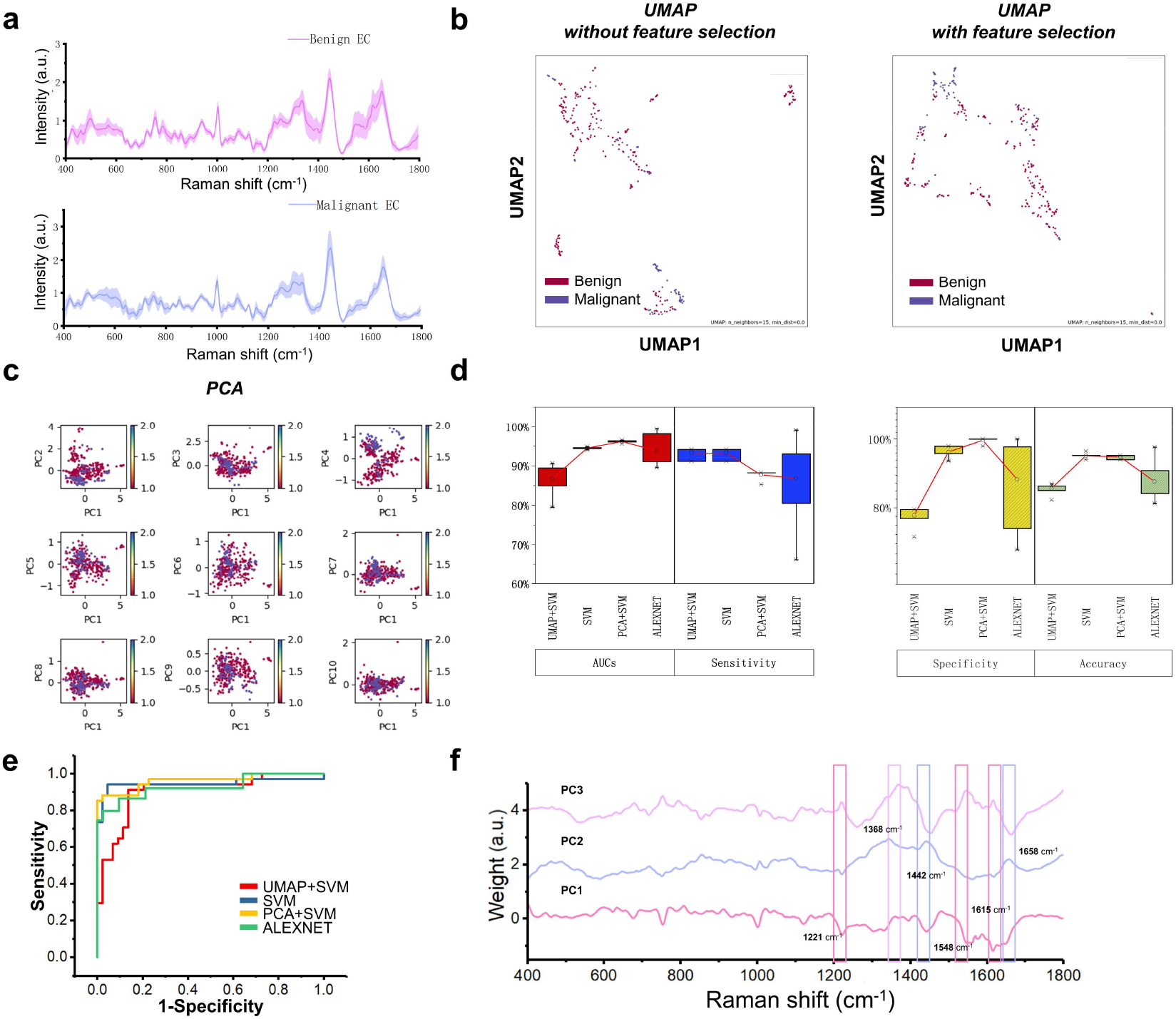
Comparison of Raman diagnostic performance using AI classification models (PCA+SVM, SVM, UMAP+SVM and AlexNet) from 80 endometrial cancer tissue sites (Begin: 40; Malignant: 40) of 20 patients. Begin and malignant endometrial tissues were differentiated based on Raman spectrum. (a) Mean raw Raman spectra of endometrial tissues ex-vivo. (b) Raman spectra differentiation using UMAP without and with feature selection. (c) Raman spectra differentiation using PCA (d) Comparisons of diagnostic confusion matrix and AUCs of endometrial cancer diagnosis by AI models. (e) ROC curve of classifications by each model. (f) Saliency curve of tumor associated biomolecules contribute to the pathological diagnosis classification by using PCA+SVM.

### Hepatoma Extracellular Vesicles (EVs) Detection

The second dataset was from fucosylated extracellular vesicles from MIHA and HepG2 cells for extracellular vesicles test with SERS spectra. The protocol for isolation of extracellular vesicles by GlyExo-Capture method refers to the manuscripts of Li et al and Chen et al^31,32^. We differentiated the cancer cells from normal cells. Here we aim to extend in vitro diagnosis (IVD) methods, previous studies demonstrated that SERS reveal logical progression biomarkers for the detection of extra-cellular vesicles in cancers diagnosis etc.^33–35^.

In **Figure 1(a)**, the sample size and divergence exist low level for cell EVs spectra, therefore, manifold learning based model may work well for EVs detection. The AUC curve shows by comparing each DL, manifold learning and ML methods, which indicates UMAP+SVM is best. By UMAP pre-process, the low dimensional projections of the Raman data were extracted, which work as input of SVM model. The low-level Raman data size and divergence fit with UMAP projection with equivalent fuzzy topological characteristics. Through UMAP preprocess, the AUC increased with 0.061 on average in ten repeats.

The average spectra show MIHA and HepG2 EVs signals in **Figure 3(a)**. We compared the visualization between UMAP and PCA in **Figure 3(b)** and **(c)**. From UMAP1 vs UMAP2 and PC1 vs PC2, we all find the two population between MIHA and HepG2 spectra. By comparing the confusion matrix of four methods of UMAP+SVM, SVM, SVM+PCA, and AlexNet in **Figure 3(d)**, we found that the best model in this case was UMAP+SVM, and the total parameter size of these two models were 0.018 MB.

**Figure 3.**
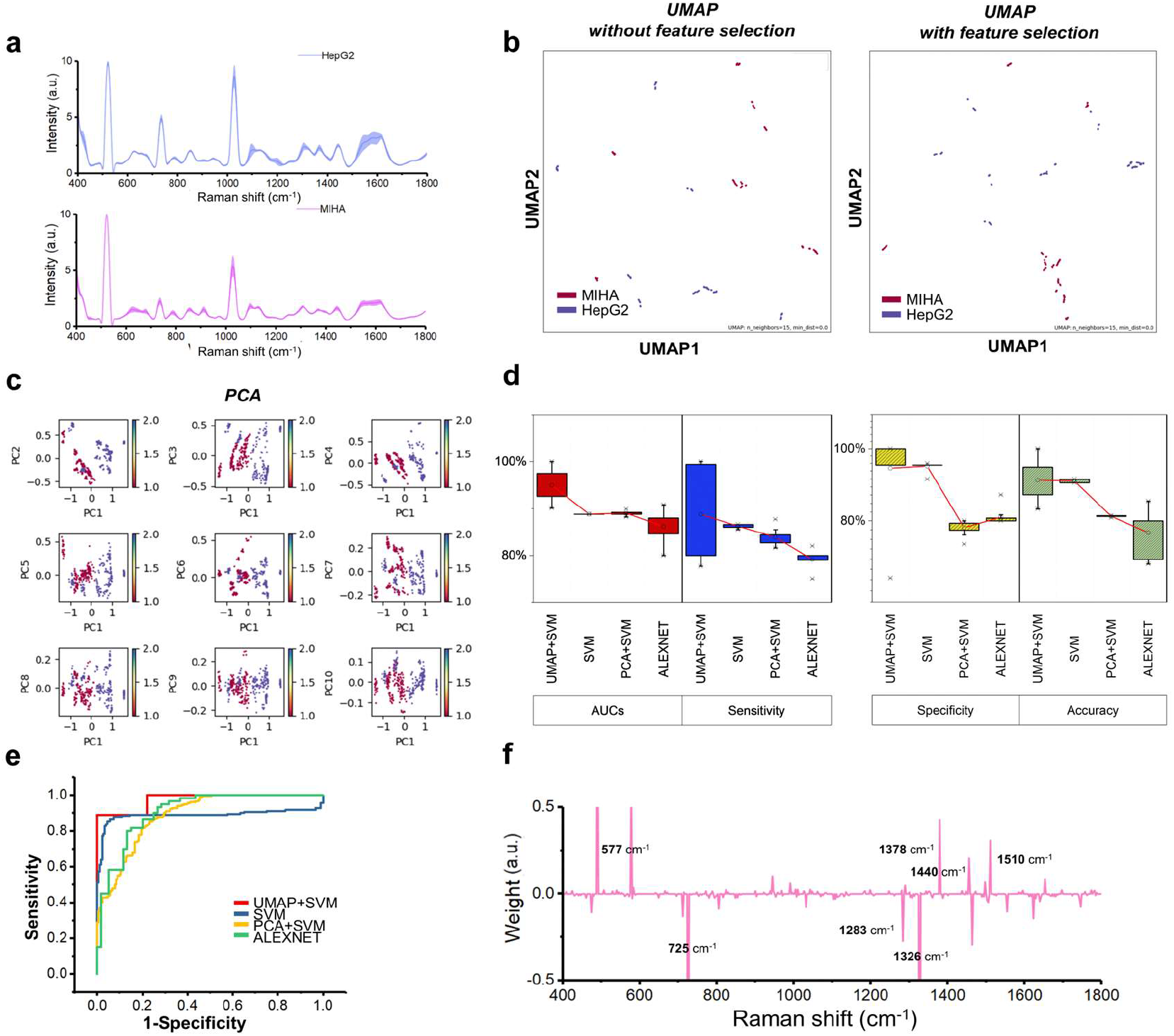
Comparison of Raman detection performance using AI classification models (PCA+SVM, SVM, UMAP+SVM and AlexNet) from 360 hepatoma cell-derived EVs sample sites (MIHA: 180; HepG2: 180) of 10 samples. MIHA and HepG2 were differentiated based on Raman spectrum. (a) Mean raw Raman spectrum of EVs extracted from MIHA and HepG2 cell line. (b) Raman spectrum differentiation using UMAP without and with feature selection. (c) Raman spectrum differentiation using PCA (d) Comparisons of diagnostic confusion matrix and AUCs of endometrial cancer diagnosis by AI models. (e) ROC curve of classifications by each model. (f) Saliency curve of Raman shift of cell type associated biomolecules contribute to the pathological diagnosis classification by UMAP.

The AUC could be 0.949±0.031 in **table S1**. Additionally, we simulated the Raman shift class weight, as shown in **Figure 3(f)**. The top contribution molecules with corresponding Raman shift were stretching mode (C=C) of carotenoid (∼1510 cm^-1^), CH_3_CH_2_ wagging and twisting of collagen, nucleic acids (∼1326 cm^-1^ and 1378 cm^-1^), unsaturated fatty acid (1283 cm^-1^), cholesterol and fatty acid (1440 cm^-1^), and symmetric breathing, phosphatidylinositol and tryptophan (∼725 cm^-1^ and 577 cm^-1^)^18,30^.

### Bacterial Identification

The third case was that we tried to identify a single bacterium for rapid antimicrobial susceptibility testing using AI assisted label-free methods. Here we collected the spectra of bacteria by Raman confocal microscopy. Previous studies demonstrated that Raman spectroscopy has the ability to achieve rapid identification of pathogenic bacteria using deep learning^3,36^. Deep learning neural networks such as a long short-term memory (LSTM)^17^ and Variational auto-encoders (VAE)^37^ have been developed to improve the accuracy of bacterial identification. We have differentiated the different six bacteria and built the Raman database.

In **Figure 1(a)**, the sample size exists high level for cell EVs spectra, therefore, DL based model may work well for EVs detection. The AUC curve shows by comparing each DL, manifold learning and ML methods, which indicates AlexNet is best. There is no significant difference with PCA/UMAP pre-process or without PCA/UMAP. This may be induced from high level divergence between categories.

The average spectra of show A. *baumannii*, E. *coli*, E. *faecium*, P. *aeruginosa*, S. *aureus* and K. *pneumoniae* bacterial signals in **Figure 4(a)**. We compared the visualization between UMAP and PCA in **Figures 4(b) and (c)**. From UMAP1 vs UMAP2, we all find the two population between melanoma cell spectra. By using the feature selection, the visualization performance improves significantly better. By comparing the confusion matrix of four methods of UMAP+SVM, SVM, SVM+PCA, and AlexNet in **Figure 4(c)**, we found that the best model in this case was AlexNet, and the total parameter size of these two models were 1.585 MB. The AUC could be 0.996±0.004 in the **table S1**. Additionally, we simulated the Raman shift class weight, as shown in **Figure 4(d)**. The top contribution molecules with corresponding Raman shift were RNA (∼710 cm^-1^), stretching mode (C-C) of proline, and CCH ring breathing of tyrosine (∼846 cm^-1^), amino acids (∼913 cm^-1^), bending mode (C-H) of phenylalanine (∼1053 cm^-1^), stretching mode (C-N) of proteins (1151 cm^-1^), and stretching mode (C-H) of tyrosine (1165 cm^-1^)^18^.

**Figure 4.**
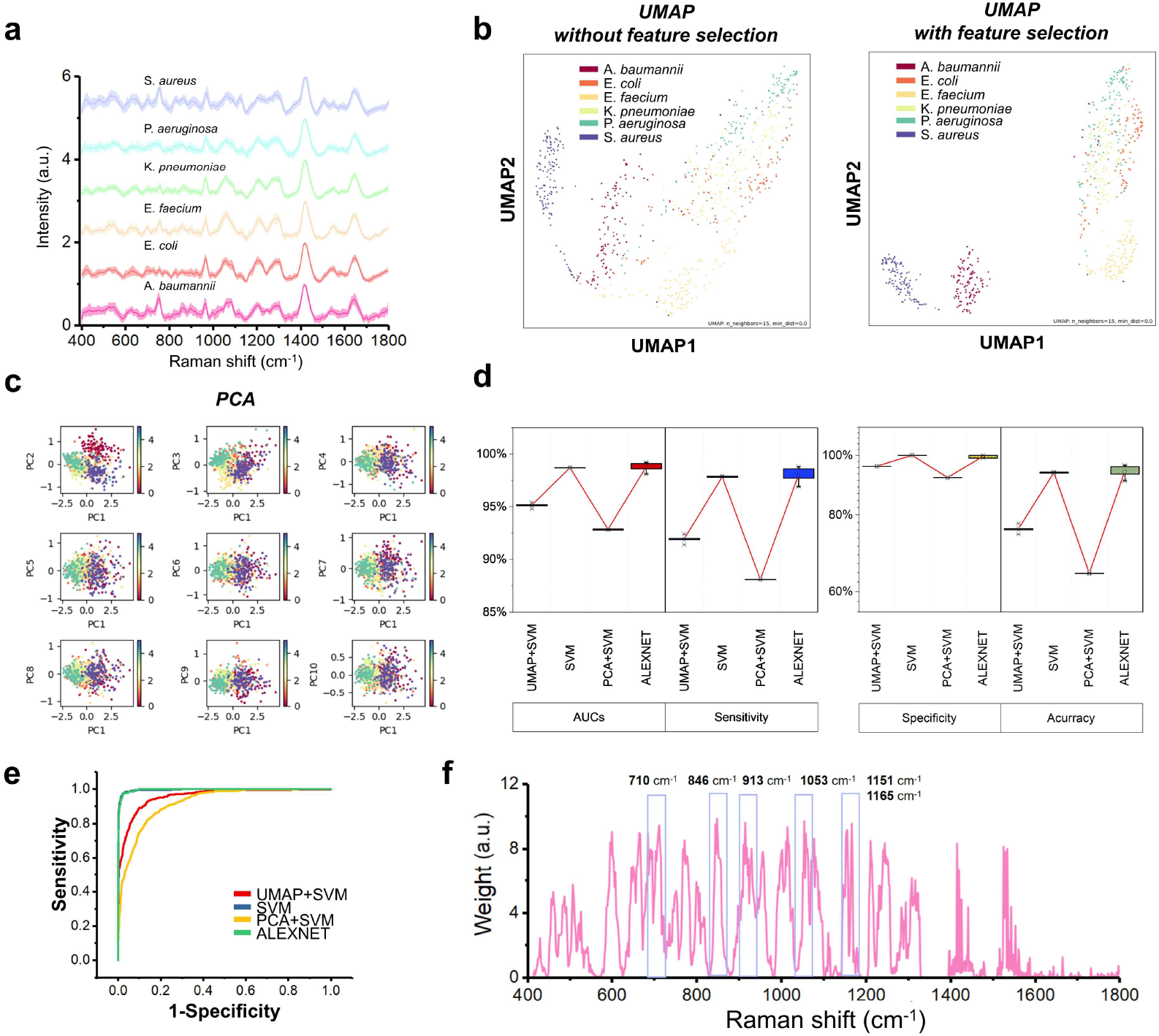
Comparison of Raman detection performance of bacterial identification using AI classification models (PCA+SVM, SVM, UMAP+SVM and AlexNet) from 720 sample sites (A. *baumannii:* 120, E. *coli:* 120, E. *faecium:* 120, P. *aeruginosa:* 120, S. *aureus:* 120 and K. *pneumoniae:* 120) of 10 patients. (a) Mean raw Raman spectrum of 6 types of bacterial ex-vivo. (b) Raman spectrum differentiation using UMAP without and with feature selection. (c) Raman spectrum differentiation using PCA (d) Comparisons of diagnostic confusion matrix and AUCs of endometrial cancer diagnosis by AI models. (e) ROC curve of classifications by each model. (f) Saliency curve of Raman shift of bacterial associated biomolecules contribute to the pathological diagnosis classification by Alexnet.

### Melanoma Cell Detection

The fourth case was that we used the public data using SERS for cancer detection in the paper^20^. Based on the public spectra, we differentiated different cell lines of melanoma, neonatal highly pigmented melanocytes with and without serum, and primary culture of normal skin fibroblasts, tumor associated fibroblasts and pure medium. In **Figure 1(a)**, the sample size and divergence exist high level, therefore, DL based model may work well for EVs detection. Comparing each DL, manifold learning and ML methods, the AUC curve indicates that AlexNet is the best, which is consistent with previous studies. There is no significant difference between classification performance with and without UMAP pre-process. This may be also induced by the high level divergence between categories.

The average spectra show the signals of cell line/cell culture with and without serum in **Figure 5(a)**. We compared the visualization between UMAP and PCA in **Figures 5(b) and 5(c)**. From UMAP1 vs UMAP2, we all find the two population between melanoma cell spectra. By comparing the confusion matrix of five methods of UMAP+SVM, SVM, SVM+PCA, AlexNet, and ResNet in **Figure 5(d)**, we found that the best model in this case was AlexNet, and the total parameter size of these two models was 6.278 MB. The AUC could be 1±0 in the **table S1**. Additionally, we simulated the Raman shift class weight, as shown in **Figure 5(f)**. The top contribution molecules with corresponding Raman shift were Fe-containing protein (1923 cm^-1^), RNA (1365 cm^-1^), amino acids, lipid (1231 cm^-1^), out-of-plane ring breathing, tyrosine (823 cm^-1^), cholesterol (609 cm^-1^), and amino acids (569 cm^-1^), which is consistent with importance analysis of previous study^18,20^.

**Figure 5.**
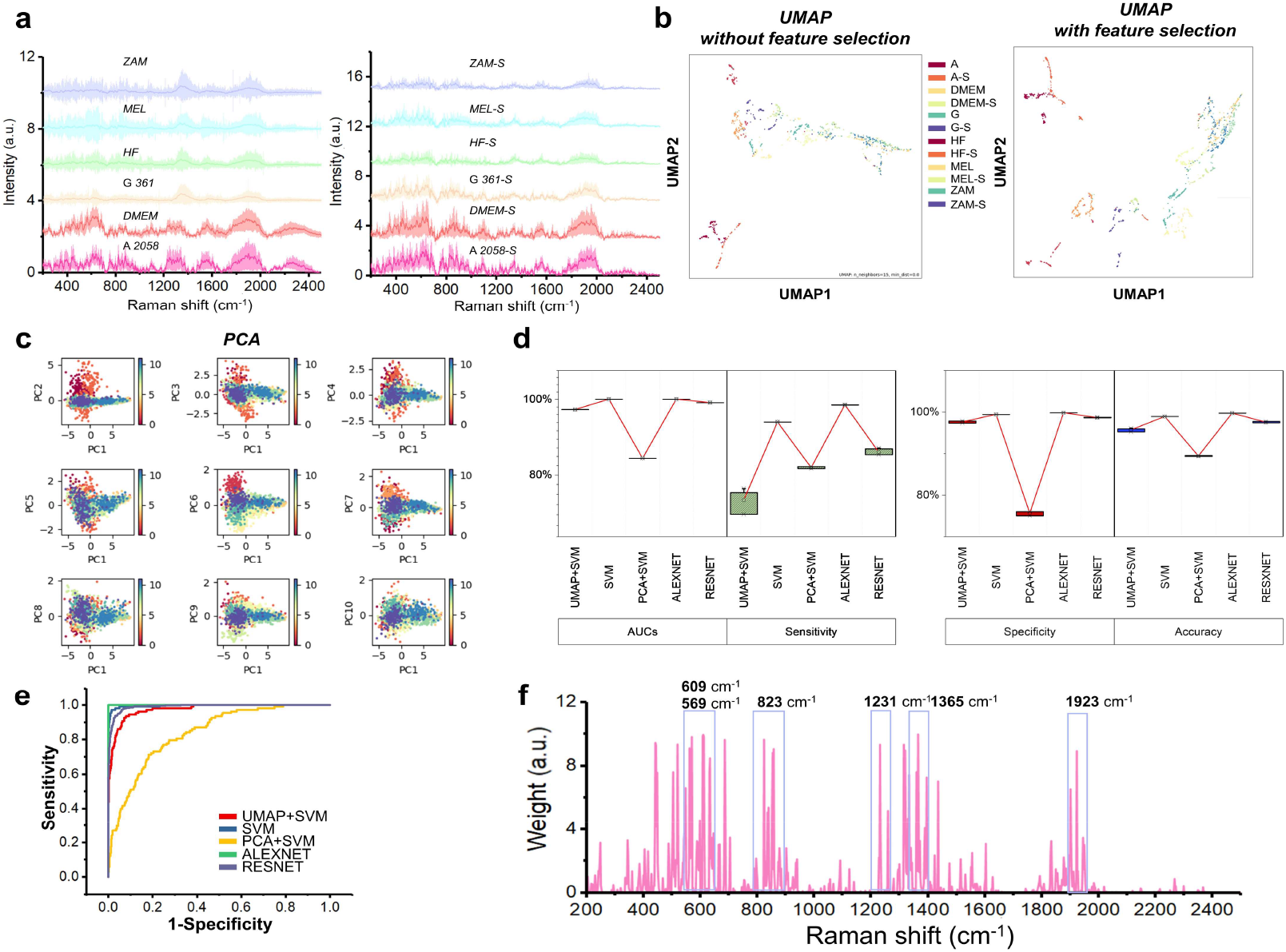
Comparison of Raman detection performance of using AI classification models (PCA+SVM, SVM, UMAP+SVM, AlexNet and ResNet) from 1881 melanoma cell sample sites (ZAM: 150, MEL: 147, HF: 168, G.361: 156, DMEM: 192, A2058: 159, ZAM-S: 147, MEL-S: 150, HF-S: 150, G.361-S: 150, DMEM-S: 159, A2058-S: 153) of 12 samples. (a) Mean raw Raman spectrum of 12 types of mela-noma cell ex-vivo. (b) Raman spectrum differentiation using UMAP without and with feature selection. (c) Raman spectrum differentiation using PCA (d) Comparisons of diagnostic confusion matrix and AUCs of endometrial cancer diagnosis by AI models. (e) ROC curve of classifications by each model. (f) Saliency curve of Raman shift of cell types associated biomolecules contribute to the pathological diagnosis classification by Alexnet.

### Diabetes Mellitus Screening

The fifth case was that we also used public in-vivo Raman spectra to demonstrate our hypothesis. Based on the public spectra in the paper^21^, we differentiate normal and Type 2 diabetes mellitus (DM2). In **Figure 1(a)**, the divergence exists high level, therefore, DL based model may work well for EVs detection. The AUC curve shows by comparing each DL, manifold learning and ML methods, which indicates AlexNet is best, which is consistent with previous study.

The average spectra show the typical signals of ear lobe, inner arm, thumb nail, median cubital vein of control and DM2 patients in the **Figure 6(a)**. We compared the visualization between UMAP and PCA **in Figures 6(b) and 6(c)**. It is still hard to differentiate control and DM2 by UMAP. By comparing the confusion matrix of four methods of UMAP+SVM, SVM, SVM+PCA, and AlexNet in **Figure 6(d)**, we found that the best model in this case was AlexNet, and the total parameter size of this model was 9.418 MB. This phenomenon is consistent with previous studies. The AUC could be 0.923±0.027 in the **Table S1**. Additionally, we simulated the Raman shift class weight, as shown in **Figure 6(e)**. The top contribution molecules with corresponding Raman shift were stretching mode (C=O) of amide I, α-helix, collagen, elastin (∼1666 cm^-1^), bending mode (CH_2_ and CH_3_) of collagen (∼1408 cm^-1^), and tryptophan, phenylalanine, RNA (∼1196 cm^-1^), and stretching mode (C-H and C-O) of lipid (1074 cm^-1^), glucose fingerprint bands (∼918 cm^-1^ and 1060 cm^-1^), and stretching mode (C-C) of glycogen, α-helix, proline, valine (∼938 cm^-1^)^18,38^.

**Figure 6.**
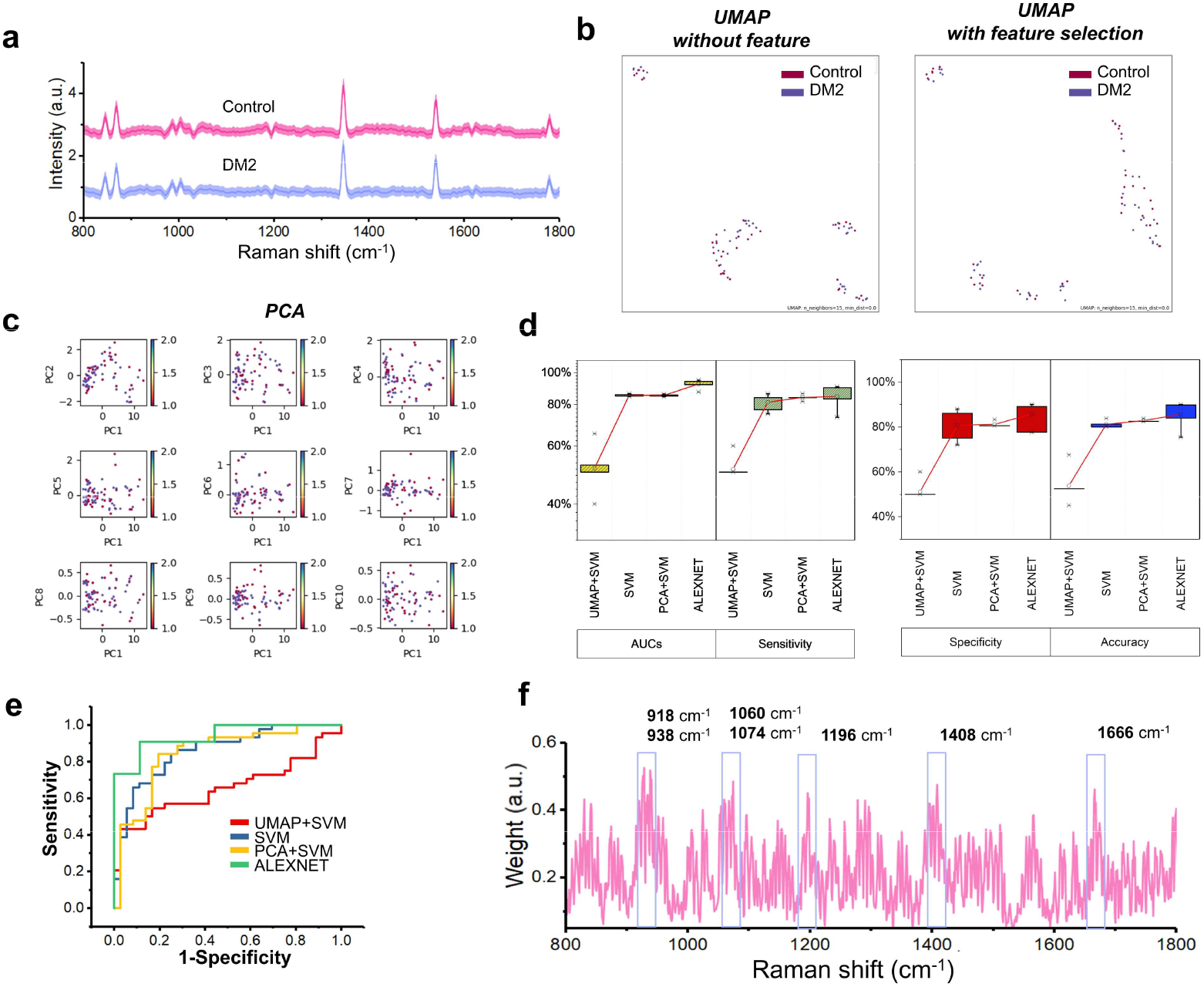
Comparison of Raman detection performance using AI models (PCA+SVM, SVM, UMAP+SVM and AlexNet) from 80 diabetes mellitus screening tissue sites (Control: 40; Malignant: 40) of 11 patients. (a) Mean raw Raman spectrum of skin tissues ex-vivo. (b) Raman spectrum differentiation using UMAP without and with feature selection. (c) Raman spectrum differentiation using PCA (d) Comparisons of diagnostic confusion matrix and AUCs of endometrial cancer diagnosis by AI models. (e) ROC curve of classifications by each model. (f) Saliency curve of Raman shift of tissue associated biomolecules contribute to the pathological diagnosis classification by Alexnet.

In this paper, we demonstrated that the parameters of best model increased as more Raman spectra size, but decreased as more KL divergence between different phenotypes. The AUC of the best model improves from 7% to 15% than others, and the best model is significantly better in confusion matrix. When developing AI classification model, we may suggest to refer to the data characteristics of spectra dataset first. This will improve the performance of Raman spectral analysis and visualization of Raman shift contributions. The input spectrum details of patient/sample number, spectral collection site number, total wave-number points, wave-number range, spectral data, significant wave points and KL divergence of five demo datasets were described in the **table S2**.

## DISCUSSION

Nowadays, selecting the model and related parameters cost a lot of time. This problem limited the clinical applications in practice by using Raman spectroscopic probe or Raman confocal microscopy in vitro and in vivo. Through studying the relation between model and Raman data, we found that the best model may be AlexNet when the data size could be more than 1MB. The best model may be ResNet when the samples source number are more than 100. The ResNet model only could be fitted when sample number and data size is all high, for instance melanoma cell detection in this paper. With the same data size, Raman data from the less sample number may match UMAP than PCA by comparing endometrial cancer diagnosis and cell-derived EVs detection. Spectra from more samples could generate enough variance as principal components. Upon five demo dataset demonstrations, we suggest that it is better to analyze the data characteristics before deciding analysis models and adjusting model parameters.

By using feature selection UMAP, the preprocess components (UMAP1 *vs* UMAP2) could be differentiated, and then analyzed for each phenotype. This will highly improve the visual performance in latent space of UMAP between different categories. We also proposed a novel method to analyze the class weight of UMAP algorithm. We simulated KL divergence decay as class weight of UMAP components with corresponding Raman shift. This class weight may help to find the Raman shift with the higher contribution to diagnosis.

## CONCLUSION

Here, we developed data characteristic assisted AI models for pathological classifications, including endometrial cancer grading, EVs detection, melanoma cell detection, bacterial identification and in-vivo diabetes mellitus screening. Through selecting AI model and adjusting model parameter (activation function, and loss function) based on data characteristics, the best classification accuracy improved around 10%, AUC improved around 0.1 respectively. All the results of these five representative Raman spectral datasets highly depend on the spectral AI classification models. According to the saliency maps, we found the classification associated biomarkers in representative datasets. For example, tryptophan, porphyrin, collagen, protein and lipids were significant molecular makers in endometrial cancer grading. In conclusion, data characteristic assisted AI classification model may improve the interpretability, robustness and accuracy of Raman spectroscopy. Such a technique will allow precise and in-time pathological diagnosis.

## Data Availability

All data produced in the present study are available upon reasonable request to the authors.

## ASSOCIATED CONTENT

### Supporting Information

That spectrum analysis algorithm scheme and mathematic methods in detail were in the **Figure S1**. The architectures of deep learning networks which we trained for Raman spectrum classifications was in the **Figure S2**. During deep-learning network training and validation processes, the loss and accuracy curves were shown in the **Figure S3**. The multi-class confusion matrix and ROCs for bacterial identification was in the **Figures S4**. The multi-class confusion matrix and ROCs for melanoma cell detection was in the **Figure S5**

## ACKNOWLEDGMENTS

This work was supported by National Natural Science Foundation of China (No. 91959120 and No. 62027824 to S. Yue), Fundamental Research Funds for the Central Universities (No. YWF-22-L-547 to S. Yue). This work was also supported by Beijing Natural Science Foundation (No.7224367 and No. L223018 to X. Chen), National Natural Science Foundation of China (No. 62205010 to X. Chen), Fundamental Research Funds for the Central Universities (No. YWF-22-L-1265 to X. Chen).

