## Supporting information for "Applications of Data Characteristic AI-assisted Raman Spectroscopy in Pathological Classification"

**Table of Contents**

**Supplementary Note S1**. Scheme of UMAP, machine learning, and deep learning algorithms for Raman and SERS spectra analysis.

**Figure S1.** Scheme of supervised model training and architectures of PCA, UMAP, and SVM models.

**Figure S2.** Architectures of AlexNet and ResNet.

**Figure S3.** The loss curve and accuracy curve during deep-learning AlexNet and ResNet training and validation processes.

**Figure S4.** Multi-class confusion matrix and ROCs for bacterial identification.

**Figure S5.** Multi-class confusion matrix and ROCs for melanoma cell detection.

**Table S1.** Sensitivity, specificity, AUC, accuracy of multivariate algorithms (N=10).

**Table S2.** Input spectra data characteristics and best model parameters.

**Supplementary Note S1**. Scheme of UMAP, machine learning, and deep learning algorithms for Raman and SERS spectra analysis.

Here we used two data reduction algorithm (PCA and UMAP) and trained three classification models (SVM, AlexNet and ResNet) for spectra subtyping. For the overall scheme of model training and test in **figure S1(a)**, we first process input Raman spectrum with or without PCA or UMAP, and we got principal components (PC1, PC2, etc.) or manifold projection values (UMAP1, UMAP2, etc.). Then we trained the classification model based on the dimensioned reduction values with labels. After training models, we predicted independent spectrum from additional samples or tissues, and tested the performance of models. Using machine learning methods of SVM, we validated the performance of models using leave-one-out cross method. Using deep learning methods of AlexNet and ResNet, we at first trained the classification models using 80% of the overall dataset. Then we tested the performance of models using the rest 20% of spectrum data. When building a spectrum analysis network, we select *linear* activation function instead of *relu* function as shown in figure. 2S. Linear activation function keeps negative correlation Raman shift with better accuracy and less loss during training networks.

The mathematic methods for PCA, UMAP and SVM were shown in the **figure S1(b)**. Using PCA, we assume that all meaningful information which contains within the variance. Through finding the maximum variance space, we could get principal components (PC1, PC2 etc.) of Raman shift with high variance. Different with PCA, UMAP used nonlinear dimensional reduction, and find a representation (UMAP1, UMAP2 etc.) of Raman data in low-dimensional space $R^N$. Firstly, a good map from Riemannian manifold $M$ to $R^N$ was found. Then Raman data $D$ is uniformly drawn from M. By simulating approximate distances in $M$ between points in $D$ that are close enough in $R^N$, we finally get UMAP values in $R^N$. Using SVM, we create a hyper plane ($\omega$*x-b=0) with minimum distance between points. SVM method only focuses on class weight from extreme points, in the meanwhile ignoring distances between other points to hyper plane. Therefore, SVM may fit well with small sample size Raman spectrum data.

The training and validation loss show in the **figure S3**, there is no over-fitting during the Raman model training. After training, we test the performance of model with the test set. The performance of multi-class confusion matrix of bacterial identification shows in the **figure S4.** For bacterial ID identification, the ROC performance of SVM and AlexNet were better than PCA+SVM and UMAP+SVM. The melanoma cell detection shows in the **figure S5**. For melanoma cell detection, the ROC performance of SVM, ResNet and AlexNet were better than PCA+SVM and UMAP+SVM. For bacterial identification and melanoma cell detection, machine learning may be enough, comparing with deep learning.

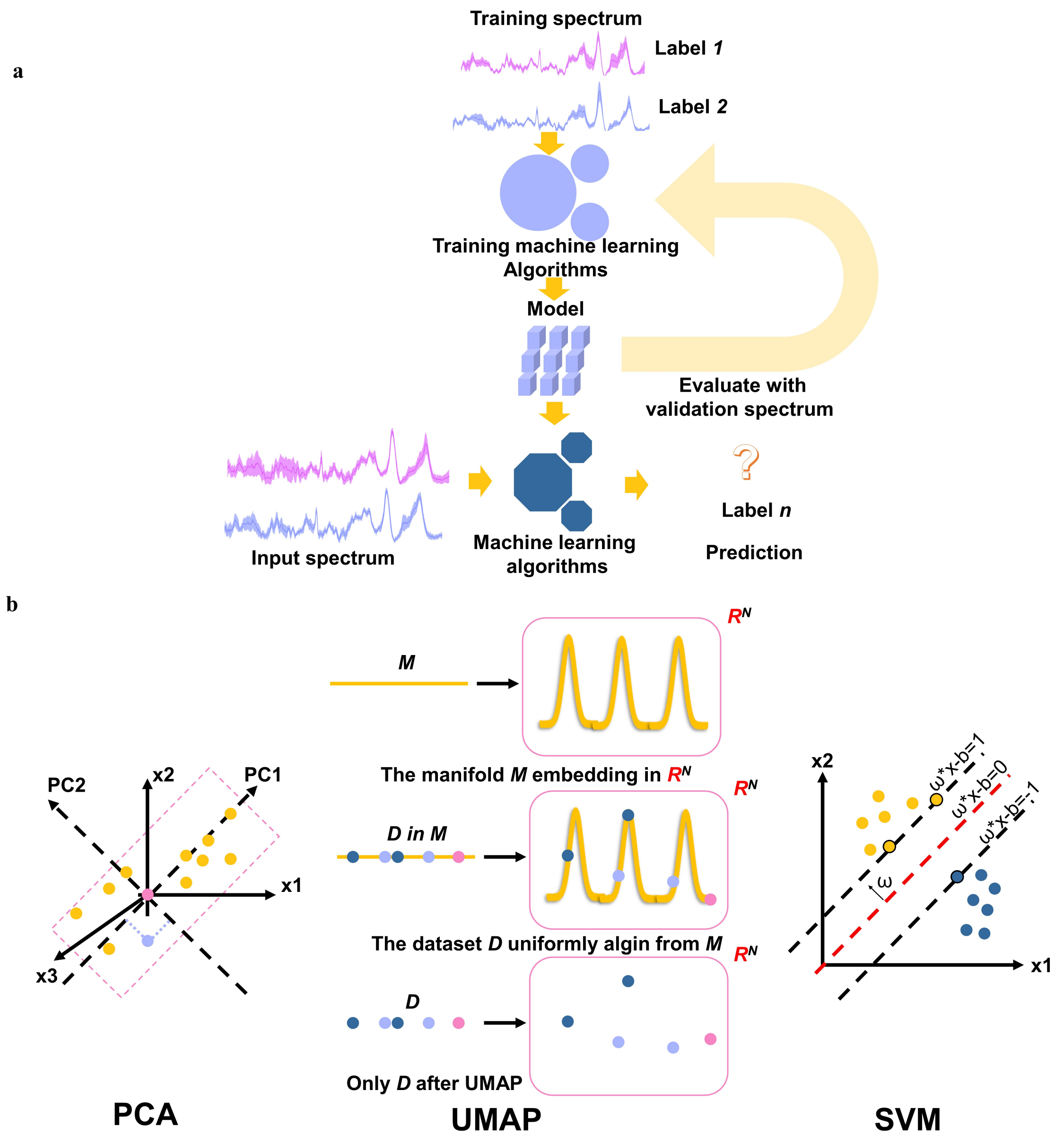


**Figure S1.** Scheme of supervised model training and architectures of PCA, UMAP, and SVM models.

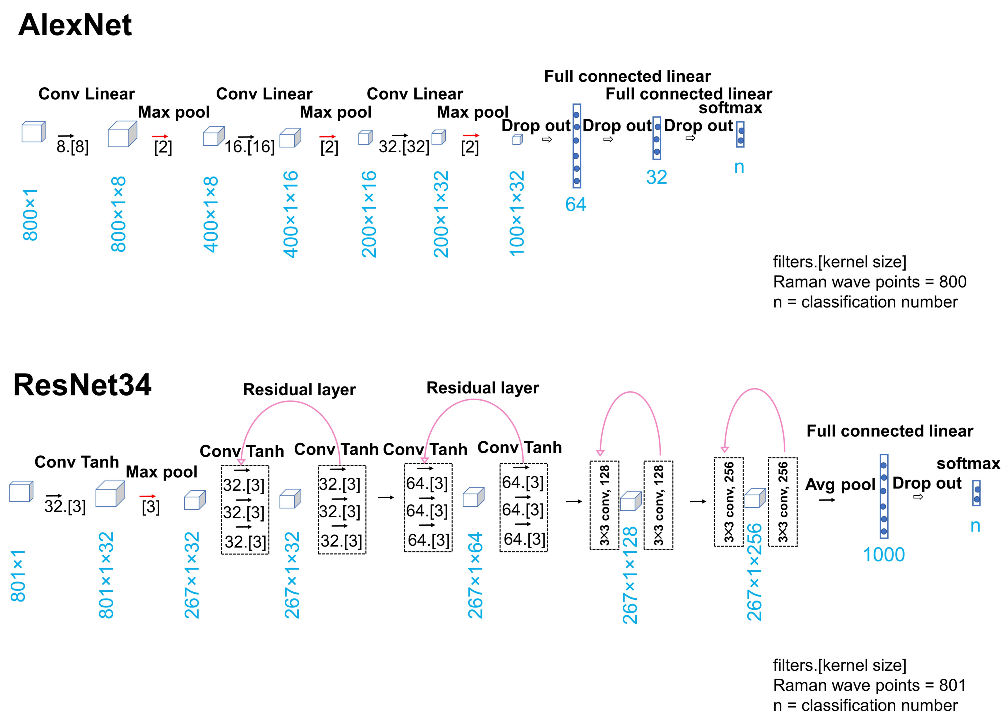


**Figure S2.** Architectures of AlexNet and ResNet.

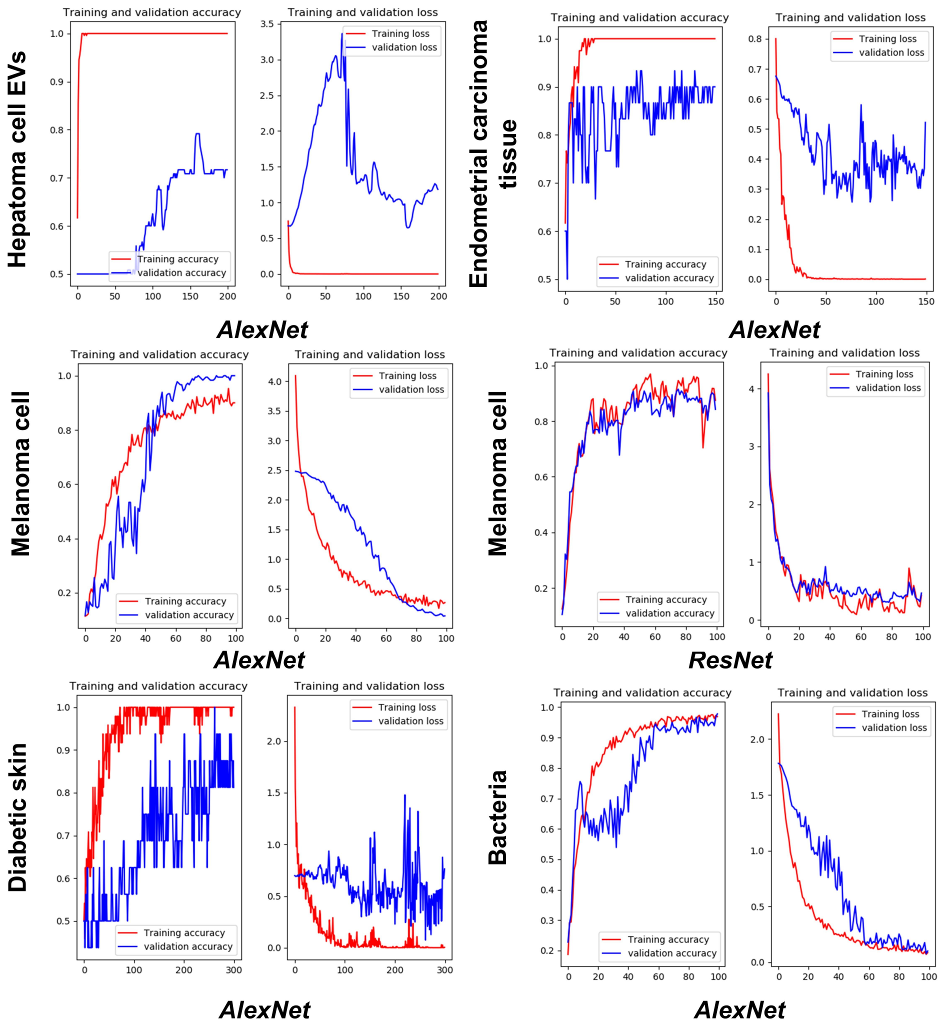


**Figure S3.** The loss curve and accuracy curve during deep-learning AlexNet and ResNet training and validation processes.

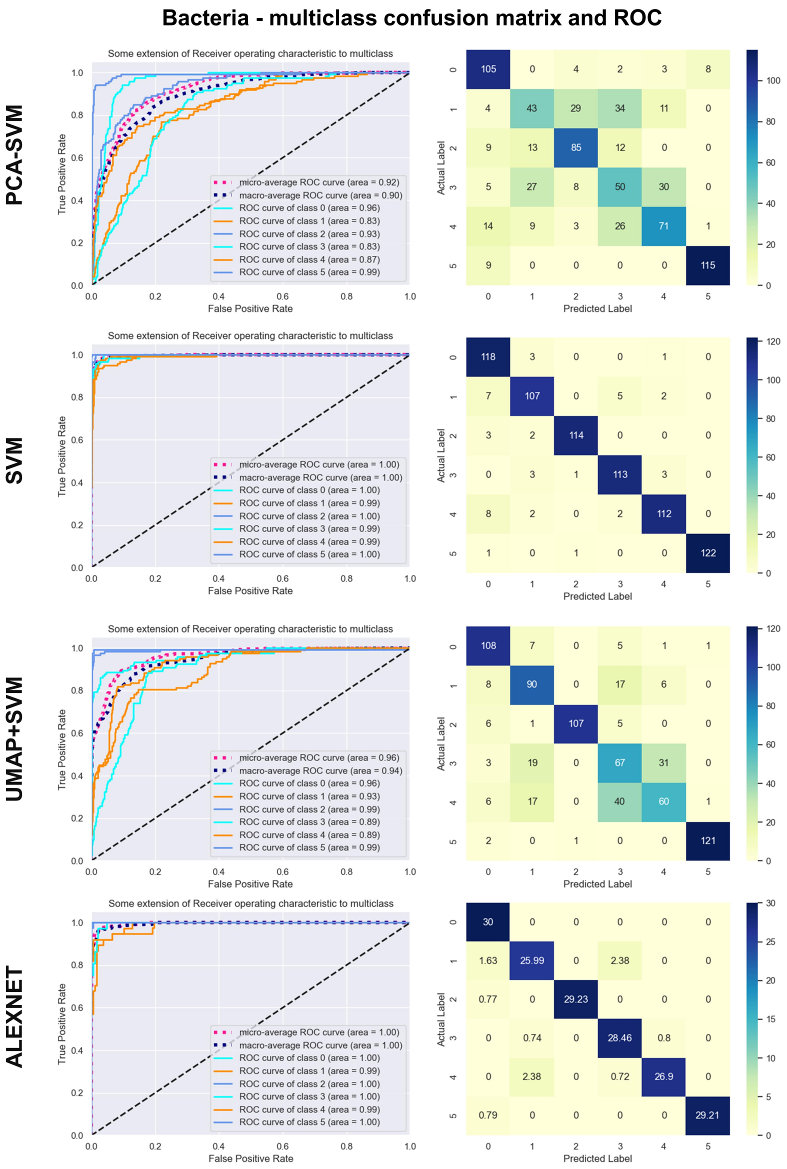


**Figure S4.** Multi-class confusion matrix and ROCs for bacterial identification.

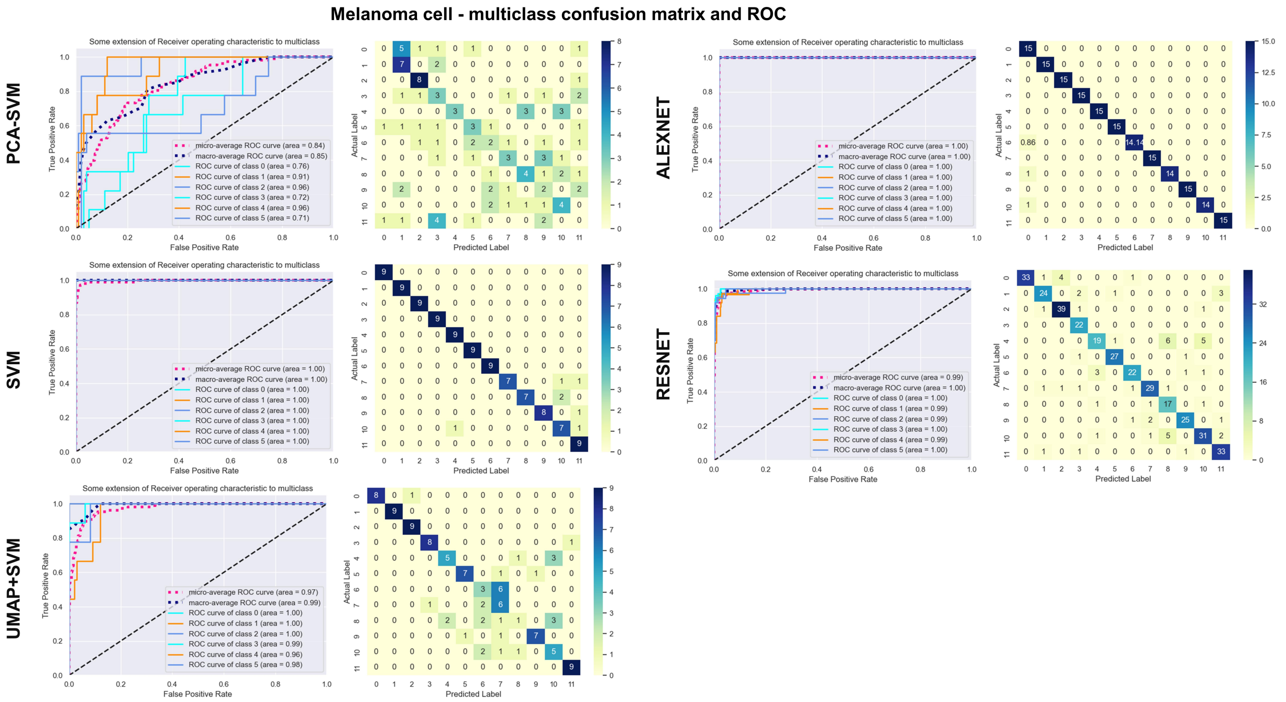


**Figure S5.** Multi-class confusion matrix and ROCs for melanoma cell detection.

**Table S1. Sensitivity, specificity, AUC, accuracy of multivariate algorithms (N=10).**

| Samples (Class categories) | Models | AUCs | Sensitivity | Specificity | Accuracy |
|---|---|---|---|---|---|
| **Endometrial carcinoma tissue (class =2)** | *UMAP+ SVM* | 0.866±0.033 | 0.932±0.014 | 0.779±0.021 | 0.851±0.012 |
| | *SVM* | 0.944±0.002 | 0.932±0.014 | 0.958±0.018 | 0.946±0.007 |
| | *PCA+ SVM* | 0.960±0.002 | 0.876±0.012 | 0.995±0.004 | 0.942±0.006 |
| | *AlexNet* | 0.937±0.038 | 0.867±0.101 | 0.876±0.116 | 0.870±0.048 |
| **Hepatoma cell EVs (class =2)** | *UMAP+ SVM* | 0.949±0.031 | 0.887±0.087 | 0.941±0.113 | 0.907±0.055 |
| | *SVM* | 0.888±0 | 0.862±0.003 | 0.947±0.013 | 0.904±0.004 |
| | *PCA+ SVM* | 0.89±0.004 | 0.838±0.017 | 0.782±0.017 | 0.811±0.001 |
| | *AlexNet* | 0.861±0.029 | 0.791±0.022 | 0.808±0.021 | 0.771±0.057 |
| **Melanoma cell (class =12)** | *UMAP+ SVM* | 0.97±0 | 0.742±0.026 | 0.976±0.002 | 0.956±0.004 |
| | *SVM* | 1±0 | 0.935±0 | 0.994±0 | 0.989±0 |
| | *PCA+ SVM* | 0.84±0 | 0.817±0.002 | 0.756±0.005 | 0.893±0.001 |
| | *AlexNet* | 1±0 | 0.983±0.001 | 0.998±0 | 0.997±0 |
| | *ResNet* | 0.99±0 | 0.855±0.008 | 0.986±0 | 0.975±0.001 |
| **Bacteria (class =6)** | *UMAP+ SVM* | 0.96±0 | 0.758±0.010 | 0.951±0.002 | 0.919±0.003 |
| | *SVM* | 1±0 | 0.937±0.001 | 0.987±0 | 0.978±0.001 |
| | *PCA+ SVM* | 0.92±0 | 0.642±0.001 | 0.928±0.001 | 0.881±0 |
| | *AlexNet* | 0.996±0.004 | 0.940±0.019 | 0.987±0.004 | 0.979±0.006 |
| **Diabetic skin (class =2)** | *UMAP+ SVM* | 0.512±0.067 | 0.51±0.031 | 0.51±0.031 | 0.537±0.060 |
| | *SVM* | 0.853±0.005 | 0.813±0.046 | 0.806±0.064 | 0.811±0.014 |
| | *PCA+ SVM* | 0.852±0.004 | 0.840±0.015 | 0.810±0.011 | 0.827±0.005 |
| | *AlexNet* | 0.923±0.027 | 0.846±0.066 | 0.856±0.055 | 0.855±0.057 |

**Table S2. Input spectra data characteristics and best model parameters.**

| Samples (Class categories) | Spectral collection site number | Patient /sample number | Total wavenumber points | Wavenumber range ($cm^{-1}$) | Spectral data (MB) |
|---|---|---|---|---|---|
| **Endometrial carcinoma tissue (class =2)** | 80 | 20 | 807 | 400-1800 | 0.655 |
| **Hepatoma cell EVs (class =2)** | 360 | 10 | 512 | 400-1800 | 0.084 |
| **Bacteria (class =6)** | 720 | 6 | 528 | 400-1800 | 9.48 |
| **Melanoma cell (class =12)** | 1881 | 12 | 2090 | 100-2500 | 70.8 |
| **Diabetic skin (class =2)** | 80 | 11 | 3139 | 800-1800 | 2.876 |
| **Samples (Class categories)** | **KL divergence (mean/max)** | **Significant wavenumber points** | **Best model** | **Parameter size of best model (MB)** | |
| **Endometrial carcinoma tissue (class =2)** | 23/189 | 114 | *PCA+ SVM* | 0.359 | |
| **Hepatoma cell EVs (class =2)** | -0.89/2.07 | 25 | *UMAP+ SVM* | 0.018 | |
| **Bacteria (class =6)** | 130/269 | 201 | *AlexNet* | 1.585 | |
| **Melanoma cell (class =12)** | 180/390 | 772 | *AlexNet* | 6.278 | |
| **Diabetic skin (class =2)** | 1.082/2.356 | 421 | *AlexNet* | 9.418 | |
